# Use of Skin Cancer Procedures, Medicare Reimbursement, and Overall Expenditures, 2012-2017

**DOI:** 10.1101/2020.05.28.20115717

**Authors:** Pranav Puri, Sujith Baliga, Mark R. Pittelkow, Puneet K. Bhullar, Aaron R. Mangold

**Affiliations:** Mayo Clinic Alix School of Medicine-Scottsdale, AZ, USA; The Ohio State University-Columbus, OH, USA; Mayo Clinic Arizona, Department of Dermatology-Scottsdale, AZ, USA

**Keywords:** Skin Cancer, Medicare Reimbursement, Utilization, Mohs Micrographic Surgery

## Abstract

The treatment of skin cancers represents a growing share of healthcare expenditures. However, little is known about how variations in payment rates relate to the use of different skin cancer procedures. This study describes recent trends in payment rates, use rates, and overall expenditures for skin cancer procedures in the Medicare Part B population. In this ecological study, we used the Medicare Physician Supplier and Other Provider Public Use File (POSPUF) to analyze trends in Medicare payment rates, use rates, and overall Medicare expenditures for skin cancer procedures from 2012 to 2017. We adjusted reimbursement rates for inflation by converting payment amounts into units of 2017 dollars. From 2012 to 2017, overall inflation-adjusted Medicare expenditure on skin cancer procedures increased 9%. Over this time period, inflation-adjusted Medicare reimbursement rates declined for each procedure class, with the exception of shave excision. Concurrently, the use rate of Mohs micrographic surgery increased 21%, while the use rate for all other skin cancer procedure classes declined. In summary, the results of this study suggest that increased use of Mohs has displaced use of other skin cancer procedures in the Medicare population.

## Introduction

Skin cancers represent the most common malignancies in the US and account for more than $8 billion of health expenditure annually.^1^ Due to an aging population, the incidence of skin cancers is increasing.^2^ In addition to topical chemotherapy, procedural treatments for skin cancers include Mohs micrographic surgery (henceforth referred to as Mohs), simple surgical excision, shave excision, as well as destructive modalities including laser surgery, electrosurgery, and cryosurgery. Medicare payments vary widely across these types of procedures.^3^ However, little is known about how variations in payment relate to the use of different skin cancer procedures. This study describes recent trends in payment rates, use rates, and overall expenditures for skin cancer procedures in the Medicare Part B population.

## Methods

We grouped procedures into the following categories using HCPCS codes: simple excision, Mohs, shave excision, and destruction of malignant lesions. Using the Medicare Physician Supplier and Other Provider Public Use File^3^, we aggregated the volume of services, number of providers, and average Medicare Part B payments from January 2012 to December 2017. We adjusted for inflation by converting payments into 2017 dollars. We averaged the number of providers for each HCPCS code within the procedure category. We define use rates as the total number of services in the procedure category divided by the year’s Medicare Part B population. This study was exempt from Mayo Clinic IRB approval. This study follows the STROBE reporting guidelines for cohort studies.

## Results

From 2012 to 2017, Mohs services had the highest average payment ($378.71), while shave excisions had the lowest ($70.99) (Table). Over this time period, payment rates declined for each procedure class with the exception of shave excision. The use rates of simple excisions, shave excisions, and destruction of malignant lesions all declined from 2012 to 2017. However, over this time period, the use rate of Mohs increased 21% from 3,554 per 100,000 Medicare beneficiaries to 4,293 per 100,000 Medicare beneficiaries (Figure). From 2012 to 2017, total expenditure on simple excisions and destruction of malignant lesions declined, while total expenditure on Mohs and shave excision increased. Total Medicare spending on skin cancer procedures grew 9% from $743,222,614 in 2012 to $806,392,161 in 2017. This was primarily driven by an $83,363,703 (18%) increase in expenditure on Mohs. Expenditure on Mohs represented 61% of overall skin cancer procedure spending in 2012 and this proportion increased to 67% in 2017.

**Table 1:** Skin Cancer Procedure Use, Payment, and Total Expenditure, 2012-2017

|  | # of Services | Number of Providers | Use Rate (per 100,000 Medicare Beneficiaries) | Average Medicare Payment | Total Medicare Expenditure | Overall Medicare Skin Cancer Procedure Expenditure | Share of Overall Medicare Skin Cancer Surgery Expenditure |
| --- | --- | --- | --- | --- | --- | --- | --- |
| **Simple Excision** |  |  |  |  |  |  |  |
| 2012 | 809,216 | 5,121 | 2,457 | \$155.32 | \$125,686,685 | \$743,222,614 | 17% |
| 2013 | 806,413 | 5,029 | 2,433 | \$152.73 | \$123,164,770 | \$765,640,334 | 16% |
| 2014 | 800,086 | 4,908 | 2,411 | \$147.24 | \$117,809,702 | \$757,342,759 | 16% |
| 2015 | 793,094 | 4,754 | 2,383 | \$147.24 | \$116,779,467 | \$792,069,533 | 15% |
| 2016 | 801,283 | 4,658 | 2,377 | \$143.98 | \$115,375,675 | \$824,099,471 | 14% |
| 2017 | 784,840 | 4,515 | 2,338 | \$137.81 | \$108,158,113 | \$806,392,161 | 13% |
| **Mohs Surgery** |  |  |  |  |  |  |  |
| 2012 | 1,170,682 | 990 | 3,554 | \$387.45 | \$453,582,606 | \$743,222,614 | 61% |
| 2013 | 1,227,926 | 1,034 | 3,704 | \$378.17 | \$464,361,335 | \$765,640,334 | 61% |
| 2014 | 1,250,876 | 1,051 | 3,769 | \$373.36 | \$467,033,139 | \$757,342,759 | 62% |
| 2015 | 1,316,146 | 1,097 | 3,954 | \$380.97 | \$501,418,473 | \$792,069,533 | 63% |
| 2016 | 1,415,383 | 1,131 | 4,198 | \$379.41 | \$537,016,751 | \$824,099,471 | 65% |
| 2017 | 1,440,886 | 1,168 | 4,293 | \$372.88 | \$537,272,681 | \$806,392,161 | 67% |
| **Shave Excision** |  |  |  |  |  |  |  |
| 2012 | 922,097 | 4,267 | 2,799 | \$59.01 | \$54,413,148 | \$743,222,614 | 7% |
| 2013 | 933,443 | 4,216 | 2,816 | \$75.12 | \$70,117,437 | \$765,640,334 | 9% |
| 2014 | 928,202 | 4,153 | 2,797 | \$73.03 | \$67,785,854 | \$757,342,759 | 9% |
| 2015 | 911,051 | 4,080 | 2,737 | \$74.63 | \$67,988,970 | \$792,069,533 | 9% |
| 2016 | 920,578 | 4,061 | 2,730 | \$73.32 | \$67,490,623 | \$824,099,471 | 8% |
| 2017 | 891,386 | 3,986 | 2,656 | \$70.83 | \$63,137,492 | \$806,392,161 | 8% |
| **Destruction of Malignant Lesion** |  |  |  |  |  |  |  |
| 2012 | 881,705 | 2,347 | 2,677 | \$124.24 | \$109,540,173 | \$743,222,614 | 15% |
| 2013 | 883,964 | 2,407 | 2,667 | \$122.18 | \$107,996,792 | \$765,640,334 | 14% |
| 2014 | 892,372 | 2,482 | 2,689 | \$117.34 | \$104,714,061 | \$757,342,759 | 14% |
| 2015 | 896,669 | 2,506 | 2,694 | \$118.08 | \$105,882,620 | \$792,069,533 | 13% |
| 2016 | 915,288 | 2,538 | 2,715 | \$113.87 | \$104,216,421 | \$824,099,471 | 13% |
| 2017 | 879,437 | 2,586 | 2,620 | \$111.23 | \$97,823,872 | \$806,392,161 | 12% |

\*\*All payment values are inflation-adjusted to 2017 dollars

**Figure 1:**
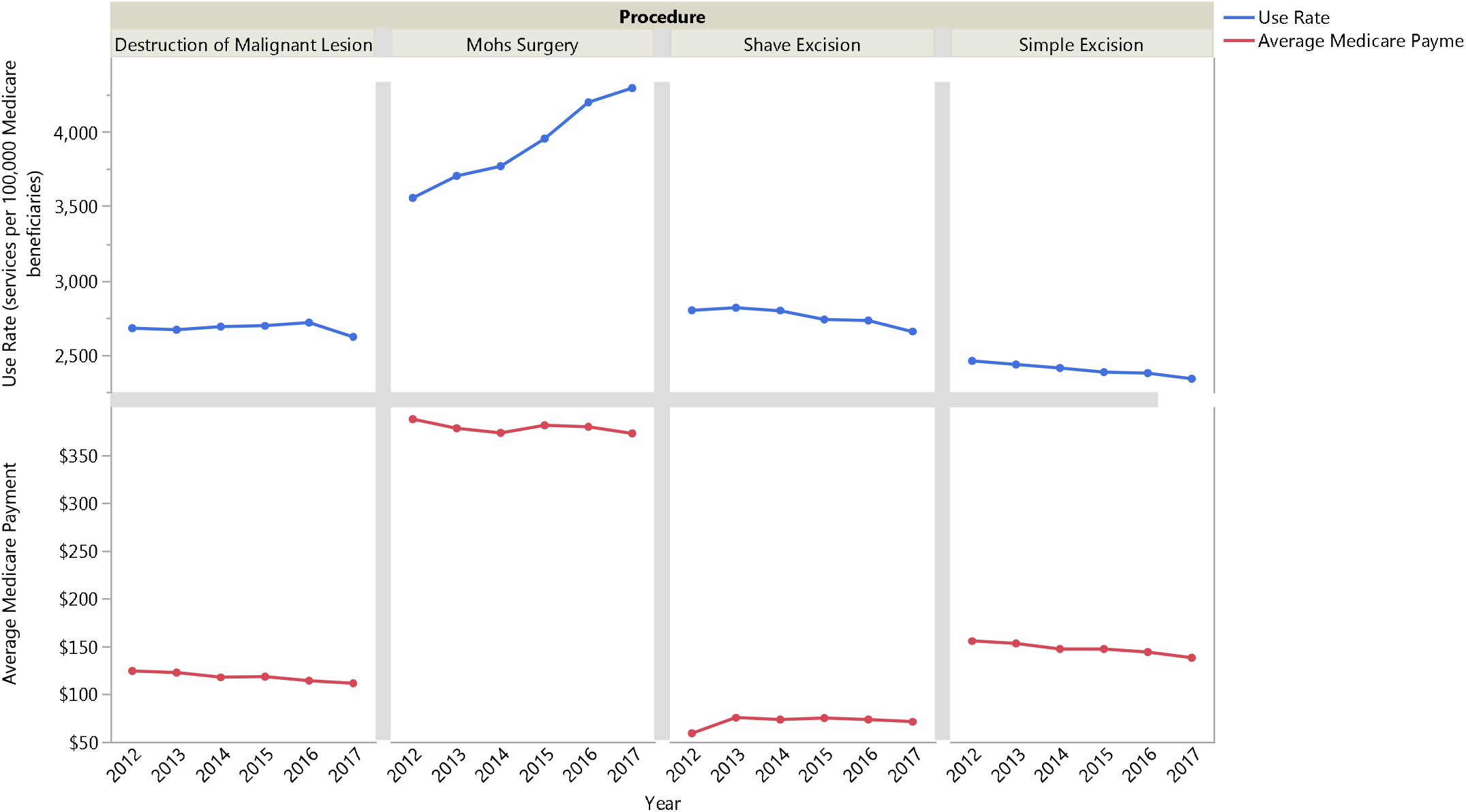
Use Rates and Payment Rates by Skin Cancer Procedure, 2012-2017

## Discussion

The results of this study suggest that increased use of Mohs has displaced use of other skin cancer procedures in the Medicare population. Medicare payment rates for Mohs are more than double those for simple excisions. Thus, from an economic perspective, Medicare fee-for-service pricing incentivizes use of Mohs in relation to other modalities.

In 2012, the American College of Mohs Surgery developed appropriate use criteria (AUC), at least in part, to reduce overuse.^4^ Yet despite the adoption of AUC, our study shows Mohs use rates have steadily increased in the Medicare population. This has driven inflation-adjusted spending growth on skin cancer procedures. Given the increasing incidence of skin cancer, further study is needed to evaluate whether the growing use of Mohs is improving value. Policymakers could potentially titrate incentives by linking payments for Mohs to adherence with AUC. Similarly, Medicare could provide a fixed, bundled payment for newly diagnosed skin cancers, thereby incentivizing more cost-effective treatments.

This study has several limitations. Since this study lacks patient-level clinical data, we cannot determine the appropriateness or outcomes of procedures. Second, the trends we describe may not generalize outside the Medicare Part B population. Third, this study did not account for ancillary costs such as repair, skin flap and pathology charges.

## Data Availability

All data is available upon request.

## References

1. Lim HW, Collins SAB, Resneck JS, et al. The burden of skin disease in the United States. J Am Acad Dermatol. May 2017;76(5):958–972.e2. doi:10.1016/j.jaad.2016.12.043

2. Collaboration GBoDC. Global, Regional, and National Cancer Incidence, Mortality, Years of Life Lost, Years Lived With Disability, and Disability-Adjusted Life-Years for 29 Cancer Groups, 1990 to 2017: A Systematic Analysis for the Global Burden of Disease Study. JAMA Oncology. 2019;5(12):1749–1768. doi:10.1001/jamaoncol.2019.2996

3. Centers for Medicare and Medicaid Services. Medicare Provider Utilization and Payment Data: Physician and Other Supplier. Accessed March 08, 2020. https://www.cms.gov/Research-Statistics-Data-and-Systems/Statistics-Trends-and-Reports/Medicare-Provider-Charge-Data/Physician-and-Other-Supplier

4. Connolly SM, Baker DR, Coldiron BM, et al. AAD/ACMS/ASDSA/ASMS 2012 appropriate use criteria for Mohs micrographic surgery: a report of the American Academy of Dermatology, American College of Mohs Surgery, American Society for Dermatologic Surgery Association, and the American Society for Mohs Surgery. J Am Acad Dermatol. Oct 2012;67(4):531–50. doi:10.1016/j.jaad.2012.06.009

